# How did governmental interventions affect the spread of COVID-19 in European countries?

**DOI:** 10.1101/2020.05.27.20114272

**Authors:** R.A.J. Post, M. Regis, Z. Zhan, E.R. van den Heuvel

## Abstract

**Background:** To reduce transmission of Coronavirus Disease 2019, European governments have implemented successive measures to encourage social distancing. However, it remained unclear how effectively measures reduced the spread of the virus, due to data complications. We examined how the effective-contact rate (ECR) among European citizens evolved over the period with implemented measures using a new data-oriented approach that is based on an extended Susceptible-Exposed-Infectious-Removed (SEIR) model.

**Methods:** Using the available data on the confirmed numbers of infections and hospitalizations, we first estimated the daily number of infectious-, exposed- and susceptible individuals and subsequently estimated the ECR with an iterative Poisson regression model, disregarding information on governmental measures. We then studied change points in the daily ECRs to the moments of the governmental measures.

**Findings:** The change points in the daily ECRs were found to align with the implementation of governmental interventions. At the end of the considered time-window, we found similar ECRs for Italy (0·29), Spain (0·24), and Germany (0·27), while the ECR in the Netherlands (0·34), Belgium (0·35) and the UK (0·37) were somewhat higher. The highest ECR was found for Sweden (0·45).

**Interpretation:** There seemed to be an immediate effect of banning events and closing schools, typically among the first measures taken by the governments. The effect of additionally closing bars and restaurants seemed limited. For most countries a somewhat delayed effect of the full lockdown was observed, and the ECR after a full lockdown was not necessarily lower than an ECR after (only) a gathering ban.

## Introduction

To reduce the transmission of Coronavirus Disease 2019 (COVID-19) European governments have implemented several non-pharmaceutical interventions aimed at reducing the number of contacts among individuals^1^. The implementation of these governmental measures differs per country, but they do follow a similar pattern. Events involving large numbers of participants are first suspended. Then the schools are being closed, shortly followed by closure of non-essential services like bars and restaurants. Finally, gathering is banned (Netherlands and Germany) or citizens are forced to stay home (United Kingdom (UK), Italy, Spain and Belgium). The latter intervention is often followed up by further restrictions, e.g. stricter surveillance by authorities. While in most countries these policies are applied to the entire nation, Italy and Spain started applying these measures in the so-called ‘red zones’ (regions where the spread started).

Stabilization of the number of daily new cases, deaths and hospitalizations was observed after the implementation of the governmental measures. These effects have been quantified in country-specific studies in which the estimated reproduction number was compared before, during and after measures were taken^2,3^. Measure-specific effects have also been estimated for some European countries^1^. These authors analyzed the observed deaths and estimated that ordering of lockdown, closure of schools, ban on public events and encouragement of social distancing would reduce the reproduction number by approximately 50%, 20%, 10% and 10%, respectively. This work assumed that the relative improvement for these interventions was the same across countries, and that measures had an immediate and constant effect (thus excluding possible delays). Another study used the number of newly confirmed infections, and showed that the effect of venue closure, gathering ban, border closure, work ban, public event ban, closure of schools and an additional lockdown reduced the reproduction number by approximately 36%, 34%, 31%, 31%, 23%, 8% and 5%, respectively. They also assumed homogeneous reductions across countries and incorporated a fixed delay of 7 days before an effect would be visible in the number of new infections^4^.

Measure-specific estimates of the reduction of the virus spread differ among the two studies^1,4^. This is not surprising, since estimation of the influence of governmental interventions is hindered by several serious limitations. The first limitation is the incompleteness of data. The number of infectious people without symptoms is largely unknown and the ‘recovery’ of non-hospitalized contagious individuals is mostly non-recorded. Both are important elements in the estimation of epidemic spread models such as the Susceptible-Exposed-Infected-Recovered (SEIR) model, to be able to determine (time-varying) contact rates and the reproduction number^5,6^. Secondly, interventions may have a partially delayed effect and it is difficult to identify how the effect of a measure changes over time. The third limitation concerns the transmission times that are commonly assumed to be exponential distributed in epidemic disease models^5^, while for COVID-19 it was shown that the incubation period is better fitted by a Weibull distribution^7–11^.

To accommodate these three limitations, we have relaxed some assumptions of the SEIR model and implemented a data-driven sequential approach to estimate time-dependent contact-rate profiles, disregarding a priori information on governmental measures. We were then able to compare the change points in the contact-rate profiles to the moments at which measures were implemented. This way, for each country separately, we observed whether the influence of measures was visible, immediate or delayed. The advantage of our country-specific analysis is that it does not require harmonization of governmental interventions across countries, it does not impose similarity across countries (e.g. decrease in the contact rate) and it allows a flexible measure-delay effect per country rather than imposing a fixed one. We illustrate our results for seven European countries that adopted different strategies.

## Methods

### Data collection

Daily counts of confirmed infections, recovered cases, and deaths were obtained from the online interactive dashboard hosted by the Johns Hopkins University^12^. These numbers were cross-validated with other sources (official sites of authorities and https://www.worldometers.info/coronavirus/). The hospitalization numbers were extracted from the official daily reports released by the public health authorities separately for each country (Appendix A). We collected data until 2020-04-09, that includes information before, during, and after governmental interventions, but does exclude relaxation of interventions. Additionally, we used data on the mobility of communities made available by Google^13^ (Appendix E).

### Epidemic disease model

We modelled the daily confirmed numbers of infections, hospitalizations, and deaths per country with an extended and time-discretized SEIR model^5,14^ (Figure 1), such that the total population, of size *N*, can be distributed over the Susceptible (S), Exposed (E), Infectious (I) and Removed (R) states (Figure 1). We assumed that the entire population is susceptible, as reported by the World Health Organization^15^.

**Figure 1.**
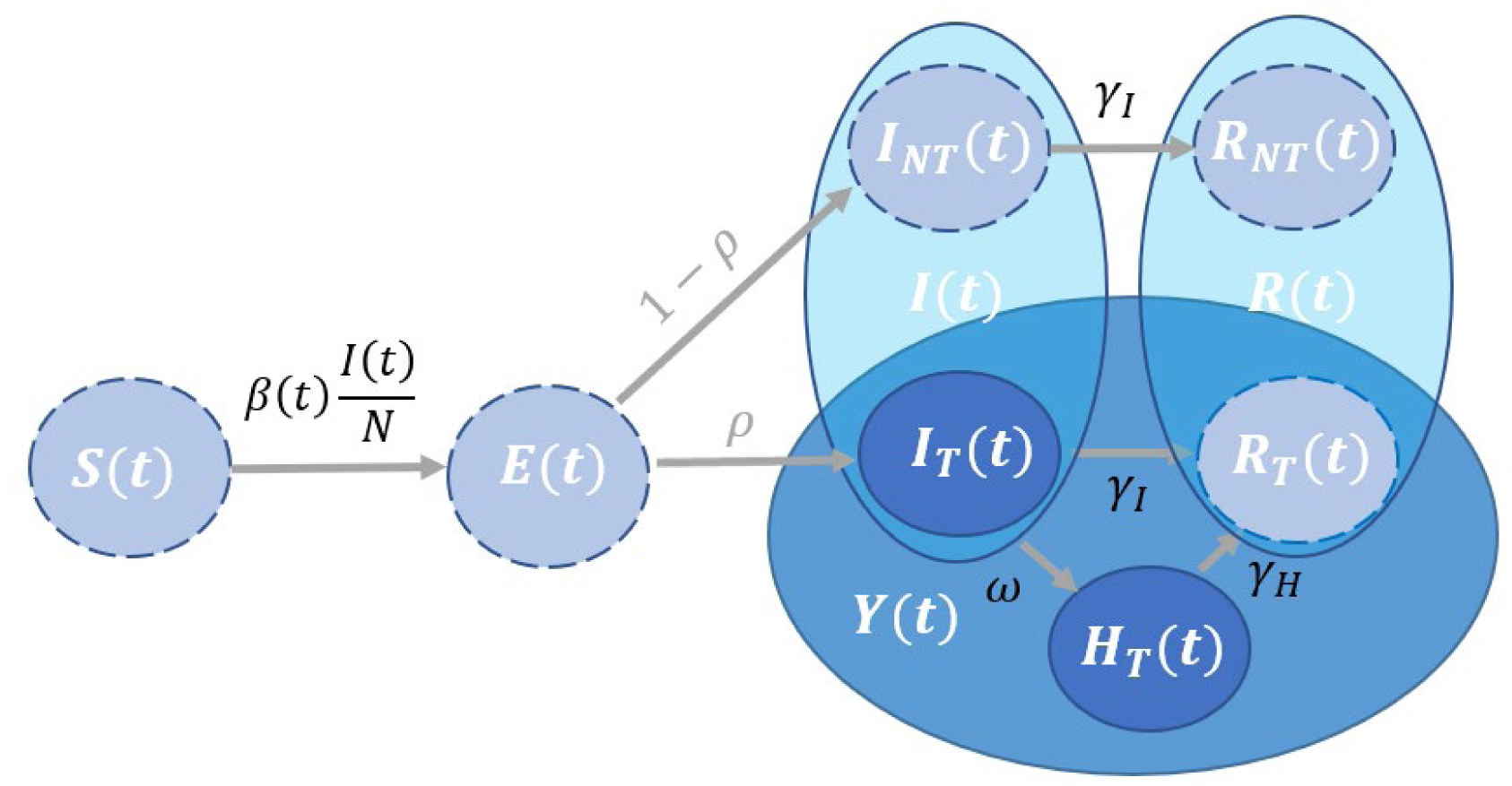
Schematic representation of the SEIR model. When there are both susceptible individuals, *S*(*t*), as well as infectious individuals, *I*(*t*), a susceptible individual gets exposed *E*(*t*) at an exponential rate of *β*(*t*)*I*(*t*)*/N*. Once exposed it takes a Weibull distributed time before the individual becomes infectious. The individual is tested with probability *ρ* and might subsequently be hospitalized. The infectious period of non-hospitalized infectious individuals is assumed to follow an Exponential distribution after which the individuals are transferred to the removed states.

A susceptible individual in *S*(*t*) can become exposed (*E*(*t*)) after an effective contact with an infectious individual. There will be an incubation period of random length before individuals become infectious and enter state *I*(*t*) (either tested *I_T_* (*t*) or non-tested *I_NT_* (*t*)). We assumed that this delay follows a Weibull distribution, with a shape parameter equal to 2·32 and a scale parameter equal to 6·50 (giving an average delay of 5·76 days), based on the preliminary study of 33 COVID-19 patients^9^. The removed state *R*(*t*)*, R*(*t*) = *R_T_*(*t*) + *R_NT_*(*t*), refers to individuals that are no longer contagious (due to death or recovery).

The number of susceptible and exposed individuals at day *t* is unknown.

The number of removed people is also only partially known, since infectious individuals are only followed-up when they are hospitalized and if they die from the disease.

Furthermore, the infectious period of infected individuals (either tested or not) is unknown, so we assumed this period to be random, following an exponential distribution with a mean, 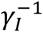, of 2·3^16,17^. For simplicity, we assumed that hospitalization takes place on the same day of appearance of symptoms, and we assumed that hospitalized individuals, *H_T_* (*t*), can no longer infect others because they are quarantined. The available information at day *t* includes the observed cumulative number of confirmed infected individuals, *Y*(*t*) *= I_T_*(*t*) *+ H_T_*(*t*) *+ R_T_*(*t*), and the daily number of new hospitalizations, ∆*H^+^*(*t*), representing individuals that transit from *I_T_* (*t*) to *H_T_* (*t*). Since the number of hospitalizations was known, we did not use the transition rates γ*_H_* and ω in this study.

The main parameter of interest is the *Effective Contact Rate* (ECR), *β*(*t*). The ECR can be interpreted as follows: The a priori probability that an infectious person upon contact meets a susceptible individual is *S*(*t*)*/N*. If we assume that every contact between an infectious and a susceptible individual leads to a transmission of the virus, then we can view *β*(*t*) as the average number of contacts on day *t*. In this case, the expected number of newly exposed individuals that one infectious person will introduce is *β*(*t*)*S*(*t*)*/N*. Since there are *I*(*t*) infectious individuals on day *t, I*(*t*)*β*(*t*)*S*(*t*)*/N* newly exposed individuals are expected on the same day. If we do not assume that all of these contacts lead to the transmission of the virus, then *β*(*t*) can be viewed as an effective number of contacts that leads to newly exposed individuals. The ECR is assumed to be time-dependent, since the introduction of different governmental interventions over time may affect the total number of contacts. Note that we did not investigate the effective reproduction number *R_e_*(*t*), which is a function of all transition rates^18,19^, since interventions only affect *β*(*t*).

### Parameter Estimation

In order to obtain an ECR profile per country, we estimated *β*(*t*) at day *t* ∊ {1,2,…, *n}*, but imposed the restriction that the ECR profile is non-increasing. We estimated the ECR from the following set of equations:

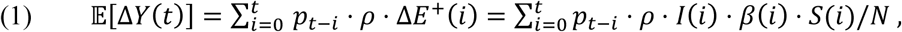

where ∆ is the symbol for daily change in the values, i.e. ∆*U*(*t*) = *U*(*t*) *− U*(*t −* 1), *E^+^*(*t*) *= E*(*t*) *+ Y*(*t*) + *I_NT_*(*t*) + *R_NT_*(*t*) and *p_t_* is the discretized Weibull probability that a susceptible individual becomes infectious *t* days after exposure. The daily number of susceptible individuals *S*(*t*) and the daily number of infectious individuals *I*(*t*) at day *t* were both unknown and derived from the observed data *Y*(*t*) (or ∆*Y*(*t*)) and ∆*H^+^*(*t*), using some assumptions on the incubation and infectious time periods (see Appendix B for details). We assumed that the number of newly exposed individuals ∆*E^+^*(*t*) at day *t* is Poisson distributed with expectation 0·5(*I*(*t*) + *I*(*t +* 1))·*β*(*t*)*·*0·5(*S*(*t*) *+ S*(*t +* 1))/*N*. Then, ∆*Y*(*t*) is also Poisson distributed with mean 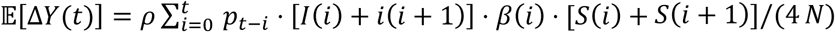. To estimate *β*(*t*), we fitted a Poisson regression model to the daily increases of tested infected individuals ∆*Y*(*t*), selecting the data from the day of the first COVID-19 fatality in the country. We assumed that *β*(*t*) did not change on the last day of our data and included *ρ* as a separate parameter in the regression analysis. Finally, we obtained confidence intervals for the estimates by means of the multivariate delta method. All estimations have been performed in R version 3.6.3.

### Sensitivity Analyses

To investigate the robustness of our analysis we conducted several sensitivity analyses: we changed the mean infectious period from 2·3 to 4·6 days^20^. We changed our mean incubation time to 6·4 days (instead of 5·76 days^9^). Additionally, we tested the influence of choosing a gamma distribution for the infectious period and an exponential distribution for the incubation time. We furthermore investigated the scenario of pre-symptomatic transmission^7,8,21^.

### Governmental Interventions

In the present study, we focused on four types of governmental interventions that were trying to limit contacts between inhabitants. Figure 2 shows the timing of these four interventions for seven European countries. A full list of governmental interventions can be found in Appendix A.

**Figure 2.**
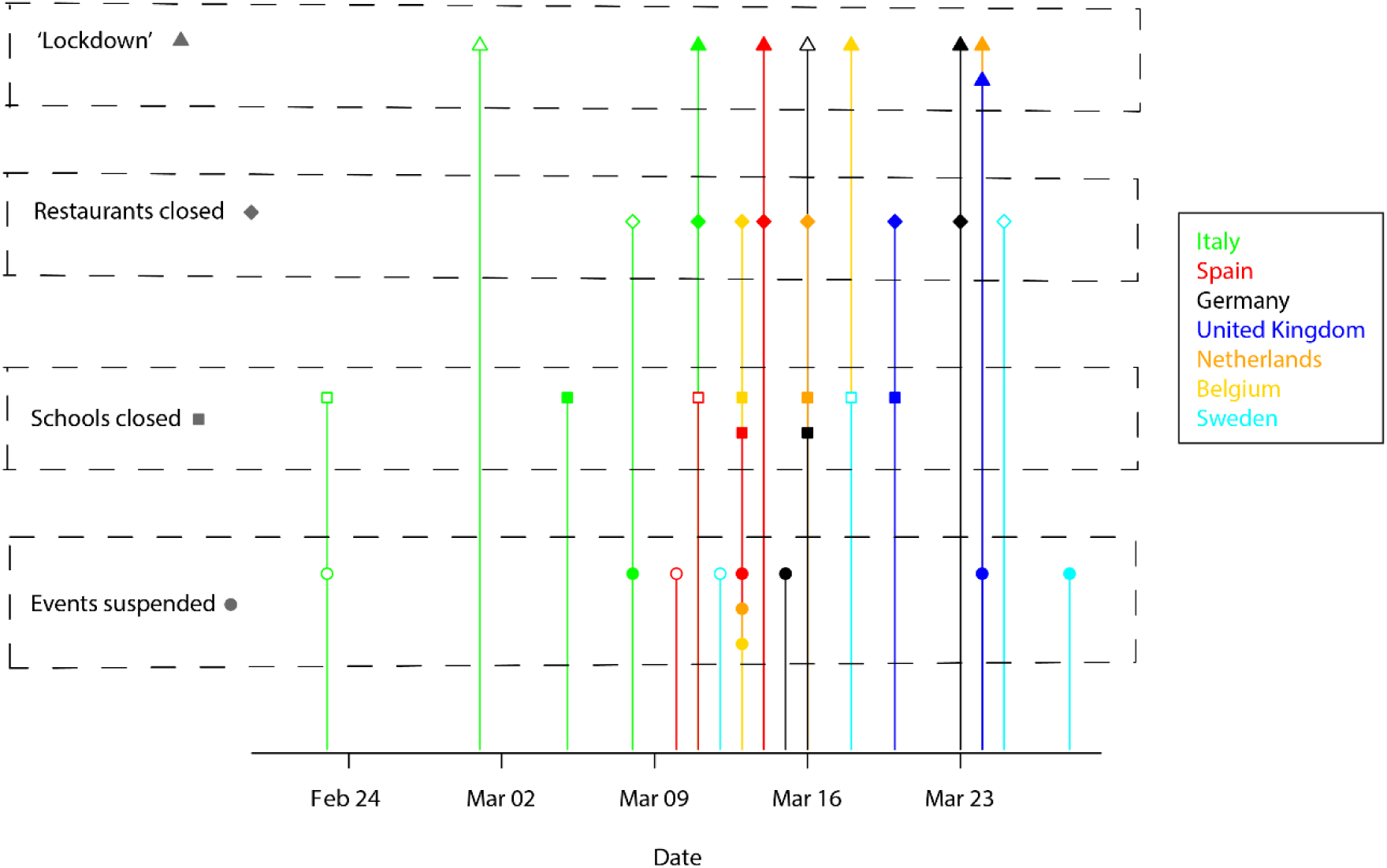
Timing of interventions. Timing of four main governmental interventions (lockdown, restaurants closed, schools closed and events suspended) for the seven European countries considered in this paper. Colors are representative for the countries, while symbols are representative for the measures taken. For each of the considered measures, we have either a filled symbol (measure taken in the whole country), or an empty symbol (measure taken in the ‘red zones’ or at a lower intensity).

## Results

The estimates of the daily ECR for each of the seven countries, together with their 95% confidence intervals, are provided in Figure 3.

**Figure 3.**
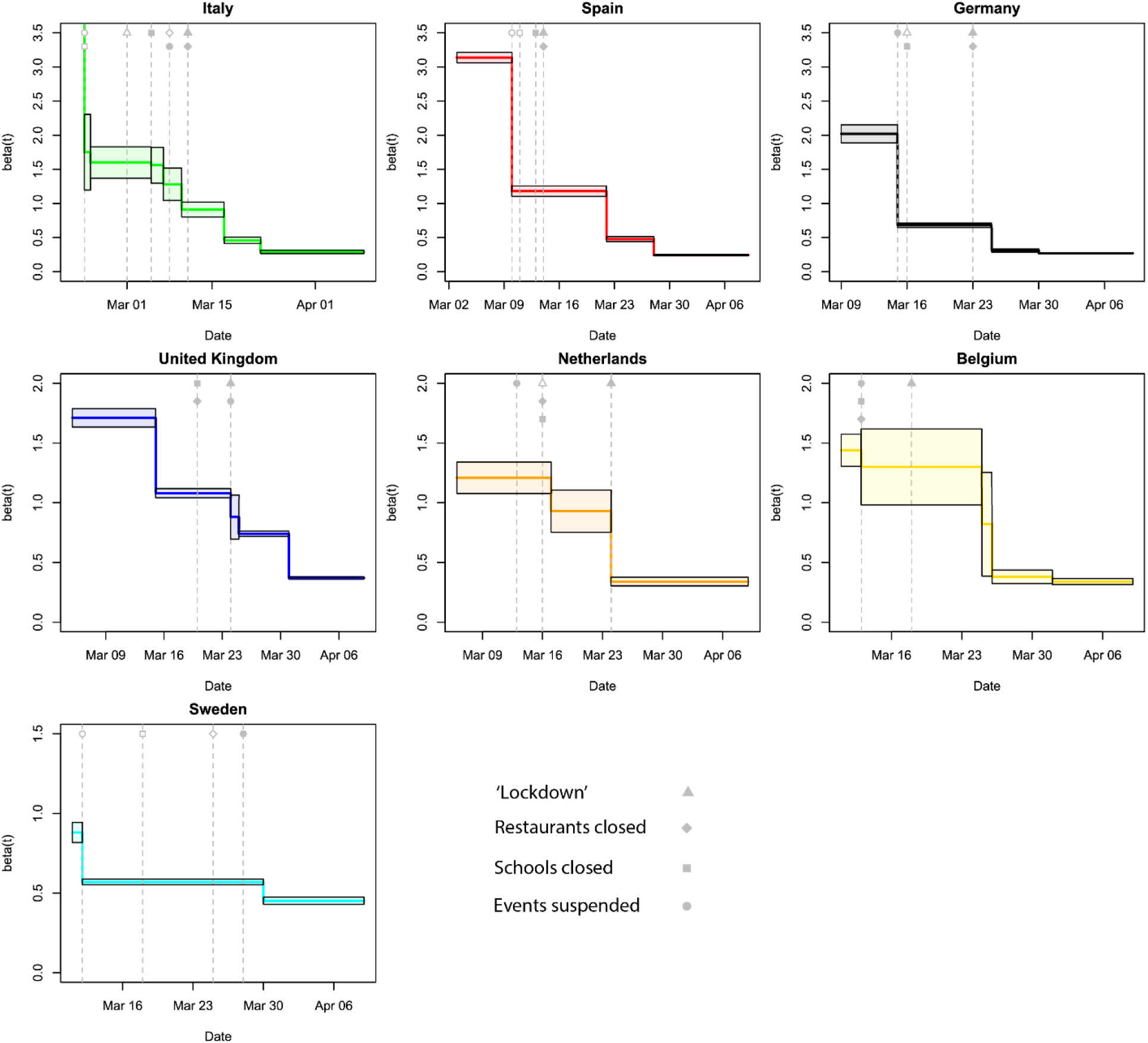
ECR profiles per country. The trend in the point estimates of the ECR 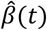 are presented as colored lines (country specific colors, following the legend in Figure 2). The black continuous lines represent the 95% confidence intervals of the point estimates. The gray vertical dashed lines indicate the moments when the governmental measures were taken, with the corresponding symbol(s) (same symbols introduced in Figure 2).

All countries have a non-increasing profile of the ECR, because of the model restriction, that consists of a few piecewise constant values. If measures had an immediate effect on the ECR, we would expect to see that the times of change points coincide exactly with at least one of the measures. However, due to the variability in the data, it is very likely that a change point is estimated just one (or two) day(s) after (or before) the real date, since daily ECRs that are similar in a period are merged to create the piece-wise constant ECRs. Then, for smaller changes in ECRs, the distance in time between the implementation of measures and the change point can become larger. In the majority of cases, we did indeed see that the drop in the ECR happens within one day of the implementation of at least one of the measures, suggesting that this measure affected the ECR, possibly in combination with measures implemented few days earlier.

Our main observations are summarized in Table 1. In Italy we observed a large decrease (9·14→1·60) after the first measures (closure of schools and event ban in the ‘red zone’) were implemented. Another serious decrease (to 1·28) occurred a day after the ‘red zone’ went into a full lockdown and events were banned in the whole country. A similar decrease (to 0·91) was found, in correspondence with the ordering of a nationwide lockdown. However, we observed a further serious decrease (to 0·46), almost a week after the lockdown was ordered. In this period, Italy enforced the lockdown with increased police forces, assisted by the army (Appendix A). For Spain, we observed two large decreases. The first change happened within a day since the banning of events and closure of schools in the ‘red zone’ (3.14→1.18). The second effect (to 0·48) became visible a week after the whole country went in lockdown. As in Italy, the Spanish government seriously increased the amount of police forces on the street days after ordering the lockdown. In Germany the first set of official measures of banning events, railway traffic reduction and closure of schools coincided with the first serious decrease in the ECR (2·02→0·68), one day before the ‘red-zone’ Bavaria went in lockdown. Two days after a full lockdown was ordered, we assessed a second change (to 0·31). For the UK, a first decrease in ECR (1·71→1·08) was visible when the UK Chief Medical Officers raised the risk-level to high (Appendix A), a measure that was initially not considered in the present study (Figure 2). A second decrease in the ECR (to 0·74) was estimated after the lockdown. The third and last decrease (to 0·37) was not traceable to the mentioned four measures but might be attributable to letters sent to 30 million households, containing details on the lockdown, rules and health information. In the Netherlands the ECR decreased (1·21→0·93) on the day after the schools and the restaurants were closed. A more serious drop (to 0·34) became visible after gathering were officially banned. In Belgium, the estimated ECR did first decrease (1·44→1·30) after events were banned and schools and restaurants were closed. More than a week after the lockdown was ordered, a serious decrease was observed (to 0·34). In Sweden, a decrease in the ECR (0·88→0·57) became visible when the Swedish government warned its citizens for the first time and banned large events. After more restrictive measures (banning of smaller events) were ordered, a small decrease (to 0·45) was estimated.

**Table 1.** Overview of the most significant changes in the ECR profiles. For each country we summarize which governmental measures were taken in close proximity to the changes estimated in the ECR 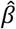. The numerical values of the 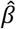 before and after the measures are presented together with the relative and absolute change.

| Country | Date | Measures | ECR ($\beta$) change | Relative change | Absolute change |
| --- | --- | --- | --- | --- | --- |
| Italy | 02-23 | Closure of schools + banning events in red zone | $9 \cdot 14 \rightarrow 1 \cdot 60$ | $0 \cdot 83$ | $7 \cdot 54$ |
| | 03-07 | Lockdown in red zone | $1 \cdot 56 \rightarrow 1 \cdot 28$ | $0 \cdot 18$ | $0 \cdot 28$ |
| | 03-10 | Full lockdown | $1 \cdot 28 \rightarrow 0 \cdot 91$ | $0 \cdot 29$ | $0 \cdot 37$ |
| | 03-17 | None (enforced police force) | $0 \cdot 91 \rightarrow 0 \cdot 46$ | $0 \cdot 49$ | $0 \cdot 45$ |
| Spain | 03-10 | Closure of schools + banning events in red zone | $3 \cdot 14 \rightarrow 1 \cdot 18$ | $0 \cdot 62$ | $1 \cdot 96$ |
| | 03-22 | None (enforced police force) | $1 \cdot 18 \rightarrow 0 \cdot 48$ | $0 \cdot 59$ | $0 \cdot 70$ |
| Germany | 03-15 | Closure of schools + event banning (+ railway reduction) | $2 \cdot 02 \rightarrow 0 \cdot 68$ | $0 \cdot 66$ | $1 \cdot 34$ |
| | 03-30 | Lockdown | $0 \cdot 68 \rightarrow 0 \cdot 31$ | $0 \cdot 54$ | $0 \cdot 37$ |
| UK | 03-15 | None (increased to high risk level) | $1 \cdot 71 \rightarrow 1 \cdot 08$ | $0 \cdot 37$ | $0 \cdot 63$ |
| | 03-24 | Lockdown | $1 \cdot 08 \rightarrow 0 \cdot 74$ | $0 \cdot 32$ | $0 \cdot 34$ |
| | 03-31 | None (information to public) | $0 \cdot 74 \rightarrow 0 \cdot 37$ | $0 \cdot 50$ | $0 \cdot 37$ |
| Netherlands | 03-17 | Closure of schools + restaurants (+ request to stay inside) | $1 \cdot 21 \rightarrow 0 \cdot 93$ | $0 \cdot 23$ | $0 \cdot 28$ |
| | 03-24 | Lockdown (fines + enforced police force) | $0 \cdot 93 \rightarrow 0 \cdot 34$ | $0 \cdot 63$ | $0 \cdot 59$ |
| Belgium | 03-13 | Closure of schools + restaurants + banning events | $1 \cdot 44 \rightarrow 1 \cdot 30$ | $0 \cdot 10$ | $0 \cdot 14$ |
| | 03-25 | None (lockdown extended) | $1 \cdot 30 \rightarrow 0 \cdot 34$ | $0 \cdot 74$ | $0 \cdot 96$ |
| Sweden | 03-12 | Banning events + warnings to public | $0 \cdot 88 \rightarrow 0 \cdot 57$ | $0 \cdot 35$ | $0 \cdot 21$ |
| | 03-29 | Stricter measures banning events | $0 \cdot 57 \rightarrow 0 \cdot 45$ | $0 \cdot 21$ | $0 \cdot 12$ |

In Table 2 we reported the estimated ECR before any interventions were implemented (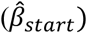), and the final ECR after all interventions were taken (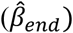). The 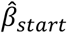 varies largely across countries, from 0·88 for Sweden to 9·03 for Italy. However, the variability in 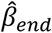 is much smaller ([0·24, 0·45]). We observed similar ECR in Italy (0·29), Spain (0·24), and Germany (0·27), while the ECR in the Netherlands (0·34), Belgium (0·35) and the UK (0·37) were somewhat higher. The ECR in Sweden (0·45), where the least rigorous measures were implemented, seemed to be the highest. The estimated fraction of tested infectious individuals *ρ*, can be found in Table 2 in Appendix D.

**Table 2.**
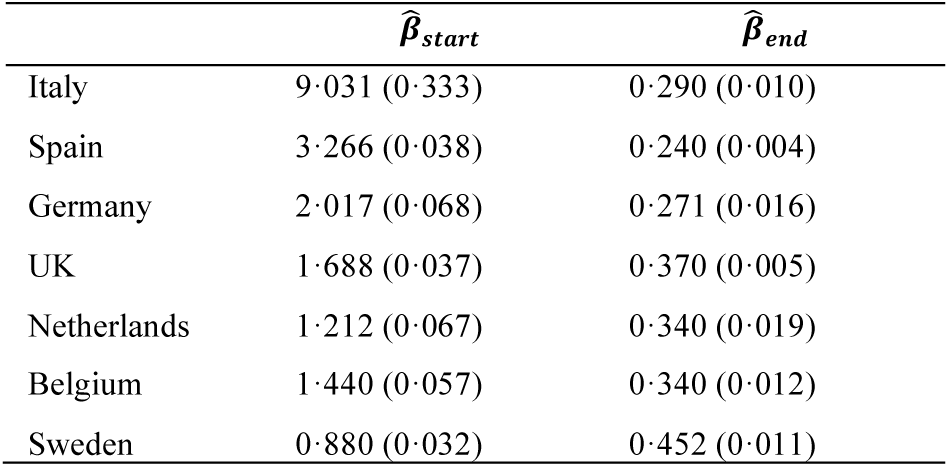
ECR change. Estimates of the ECR at the start of the considered time window, 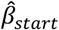, and the rate at the end of the window 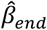 with corresponding standard errors.

In Figure 4, the daily observed number of new confirmed infections ∆*Y*(*t*) (the points in the graph) are shown together with the fit of our Poisson regression model (the continuous lines) and their corresponding 95% prediction intervals. The results of the sensitivity analyses can be found in Appendix C, demonstrating that our ECR profile was robust against violation of our assumptions.

**Figure 4.**
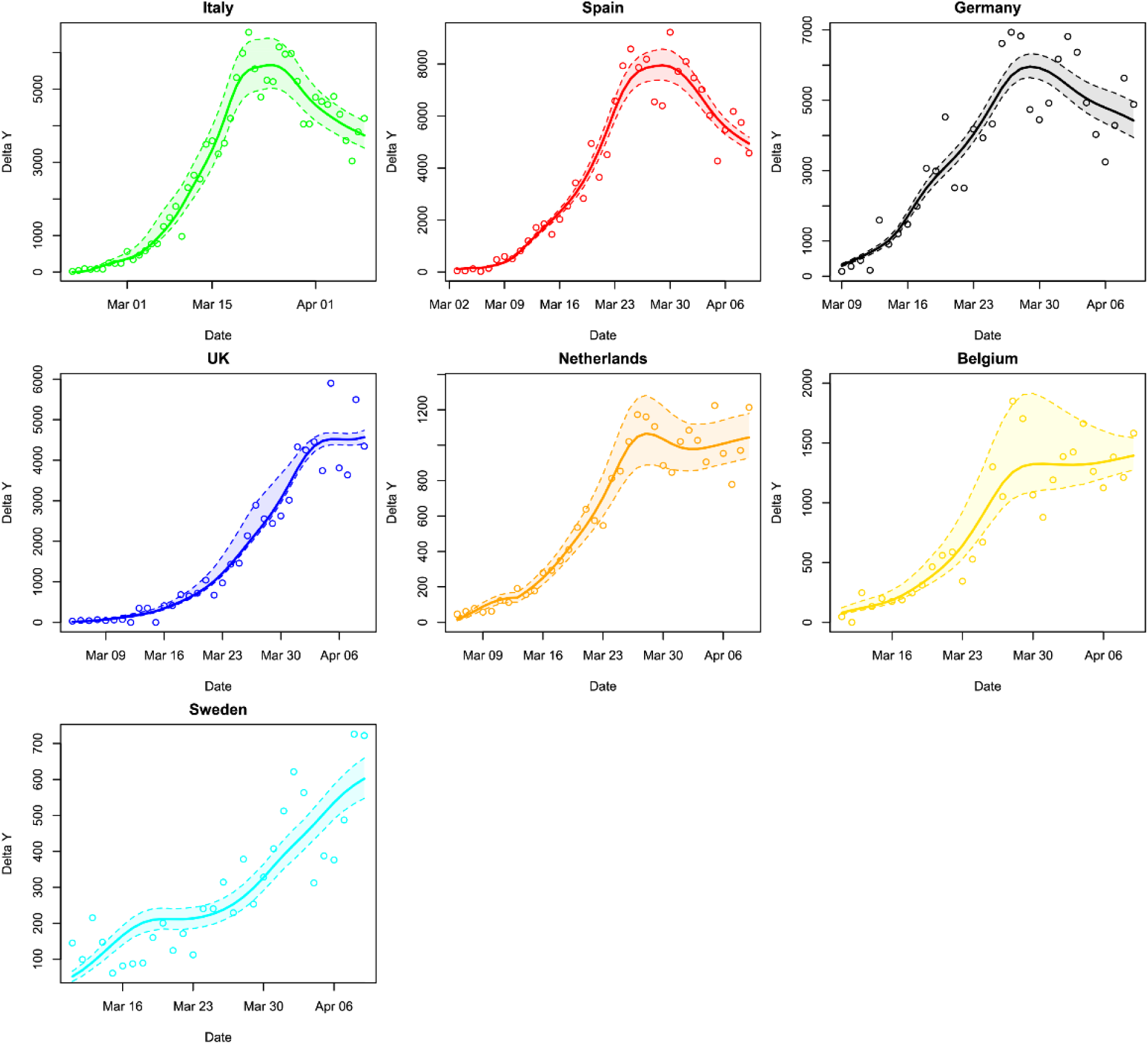
Observed and estimated number of daily new confirmed infections. The daily counts ∆Y(t) (points) are presented together with the estimated expected daily new counts from our Poisson regression model (continuous line). Colors are again representative of countries (following the legend in Figure 2). Furthermore, 95% prediction intervals are presented based on 10000 simulations assuming multivariate normality of the maximum-likelihood estimates of 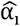.

## Discussion

In this study we have extended the SEIR model to allow for general transition-time distributions and developed an estimation strategy in which we first retrieved the latent number of infectious and susceptible individuals per day, and subsequently estimated the ECR per day. This way, we did not make a priori assumptions on consistency across countries and delays in the effect of governmental interventions. Our findings show that change points in the daily ECRs overlap with moments of governmental interventions. Overall, the first set of measures taken by the governments seems to have an immediate (although heterogeneous) effect. This first set of measures typically included the closure of schools and daycares, as well as banning events. Implementing a full lockdown after the first set of measures, either locally or nationally, seemed to coincide with changes in the ECR, but in several countries only when this was enforced with increased police surveillance.

Although the effect of closing schools and banning events has been demonstrated in previous studies^1,4^, our findings suggested a stronger (combined) effect of the first set of measures than found before. On the contrary, these studies observed more serious effects of a gathering bans, lockdowns, and closure of bars and restaurants^1,4^. These differences may be explained by the additional model restrictions (constant contact rates, country homogeneous effect sizes, and fixed delay in the effects) that these studies implemented. Therefore, they could not determine whether the effect of the full lockdown did change over time as we observed. Our observation is in line with the Google mobility data (Appendix E) that suggests that mobility did further reduce in the days after the ordering of the lockdown in Italy, Spain and (less) in the UK and Belgium. The closure of bars and restaurants frequently coincided with the moment of lockdown (Italy, Spain and Germany) or with closing schools (UK, Netherlands, Belgium), thus it is difficult to distinguish the effect of the different measures. In Italy and Spain, the combined measures had only limited influence on the contact rate, and in Sweden national restrictions on restaurant and bars had no effect. So, it seems unlikely that the closure of bars and restaurants had an additional effect (to the previous measures taken) on the ECR.

Our findings strongly suggest that the ECRs in Italy, Spain, and Germany were very similar after all measures were implemented. This was unexpected since more restrictive measures were taken in Italy and Spain, but it was consistent with other findings^1^. The final ECRs for the Netherlands, Belgium and the UK were also comparable but somewhat higher. The difference between the two groups was not the result of lower compliance to lockdown restrictions, since the Google mobility data showed similar decreases in activity in Belgium and the UK as observed in Spain and Italy. We did therefore conclude that gathering bans (as ordered in Germany and the Netherlands) were as effective as full lockdowns (as implemented in Italy, Spain, UK and Belgium). An ECR for COVID-19 of 0·30 seems to characterize a society in which citizens only have interactions for essential needs (e.g. grocery shopping and travel of healthcare workers). It is important to observe that despite the fact that the ECRs of the countries are comparable after all the interventions, the ECRs did seriously differ in the period before interventions were implemented. This suggests that the different interventions cannot have the same (absolute and relative) effect across countries, as assumed in previous studies^1,4^.

Several limitations of our study need to be addressed. First, our method did not provide estimates of effect sizes for the different measures, contrarily to what is done in previous papers^1,4^, and therefore our results may seem more exploratory. We explicitly made this choice, since interventions were often taken simultaneously and there was no information about the delay on the effect of various measures. All measures try to directly lower the ECR, but the success highly depends on the compliance of the citizens. This was supported by the Google data, showing that reductions in mobility do not align directly with implemented measures. A second limitation of our approach is the implemented restriction, imposing non-increasing ECRs. This choice was motivated to avoid additional oscillations in the parameter estimate due to the large variability present in the data (shown by the observations outside the prediction intervals in Figure 4), possibly a result of delays in testing and reporting. Thirdly, our analysis was based on a few assumptions (although supported by other research), like the choice of distributions for the incubation and infectious periods. The influence of our modelling assumptions was investigated in the sensitivity analysis (Appendix C) and showed that variation in the shape of ECR profile was limited for all countries and did not shift the change points. Lastly, we assumed that the fraction of tested infectious individuals *ρ* was unknown but constant, while it may change with test policies within countries. Again, the sensitivity analysis showed that the percentage of tested individuals did not influence the shape of the ECR profile.

The main strength of our work is the data-oriented approach: we focused on the information present in the data and modelled it with an extended version of a well-known epidemic disease model, based on a limited number of assumptions. The data demonstrated essential change points in ECR and the sensitivity analysis demonstrated that our assumptions have little influence, making the ECR profile trustworthy. This solid ECR profile could then be connected to moments at which governments implemented measures, evaluating direct and delayed effects. We believe that such approaches are more valuable than the approaches used so far, since the data collection does not satisfy assumptions that are normally valid in other areas of epidemiological research. With our approach and the data of multiple countries, we have been able to disentangle the influence of several governmental interventions. We have showed an immediate influence of banning events and closing schools on the spread of the virus, and a somewhat delayed effect of the full lockdown. Closing bars and restaurants seems to have only a limited effect instead.

Although we observed that the first set of governmental interventions had an immediate causal effect, the causal pathway of the first measures remains unclear. This set of measures might have made citizens realize the severity of the situation, which consequently made them reduce their social activity. This would confound the effect estimates of the physical constraints introduced by the interventions and should thus not be overlooked. When governments will decide to relax their interventions, the ECR might increase both as a direct result of the vanishing physical restrictions, and indirectly via the relaxation in the behavior of citizens. As a result, the ECR before the intervention was implemented could seriously differ from the ECR after the same intervention is lifted again. Prediction studies for exit strategies presenting counterfactual scenarios^1,22–24^ should therefore be dealt with caution.

## Data Availability

Data used in this study is obtained from publicly available sources that are referred to in the main text and appendices.

## Contributors

RAJP designed and programmed the model, performed the statistical analyses and made the Figures. ERH supervised the research. ZZ and MR collected and verified the data of daily cases and hospitalization. All authors interpreted the results, contributed to writing the article, and approved the final version for submission.

